# Cortical Thinning in Adults with Prader–Willi Syndrome: Distribution Across Functional Networks and Associations with BMI and the GH/IGF-1 Axis

**DOI:** 10.64898/2026.11.04.26367090

**Authors:** Alex Moreno-Armengol, Laura Blanco-Hinojo, Laia Casamitjana, Olga Giménez-Palop, Jesús Cobo, Jesús Pujol, Gerard Martínez-Vilavella, Joan Deus, Susanna Esteba-Castillo, Rocío Pareja, Assumpta Caixàs, Cleofé Peña-Gómez

## Abstract

Prader–Willi syndrome (PWS) is a rare neurogenetic disorder associated with neuropsychiatric, metabolic, endocrine, and physical abnormalities. Although previous neuroimaging studies have identified regional gray matter alterations, cortical thickness (CT) abnormalities and their distribution across large-scale functional brain networks remain poorly characterized in adults with PWS. We investigated cortical morphology in 23 adults with PWS and 22 age- and sex-matched healthy controls who underwent structural MRI. Cortical reconstruction and surface-based morphometric analyses were performed using FreeSurfer. Between-group differences in CT were assessed using age, sex, and intracranial volume as covariates. Associations between CT and clinical, cognitive, anthropometric, and endocrine measures were examined using Spearman correlations. Significant cortical regions were further mapped onto an a priori functional network atlas. Adults with PWS showed reduced CT in a left-hemispheric cluster involving the inferior precentral gyrus, insula, inferior temporal cortex, and orbitofrontal cortex. This cluster overlapped predominantly with the somatomotor, auditory, cingulo-opercular, language, and frontoparietal networks. Within the PWS group, lower CT was associated with higher body mass index and poorer global functioning. CT was positively associated with IGF-1 levels and was greater in participants with a history of growth hormone therapy during childhood. These findings indicate cortical thinning across regions belonging to multiple functional networks involved in sensorimotor, attentional, language, and executive processes, extending previous volumetric findings. The associations with body mass index, functional measures, and the GH/IGF-1 axis suggest that cortical morphology may reflect an interplay between genetic vulnerability and potentially modifiable metabolic and endocrine factors, raising the possibility of a neuroprotective role for GH/IGF-1 signaling.

## Introduction

Prader–Willi syndrome (PWS) is a rare neurogenetic disorder caused by loss of paternal gene expression in the imprinted 15q11–q13 region, typically resulting from paternal deletion, maternal uniparental disomy, or an imprinting defect [1–3]. PWS affects approximately 1 in 10,000–30,000 individuals [1,2] and is associated with substantial health and functional burden, including increased healthcare utilization, reduced life expectancy, and high levels of disability and dependence. The syndrome is characterized by a distinctive developmental course [4]. During infancy, affected individuals typically present with severe hypotonia and feeding difficulties, whereas later childhood and adulthood are characterized by hyperphagia, obesity in the absence of strict environmental control of food intake, developmental delay, intellectual disability, behavioral disturbances, and endocrine dysfunction [1,5,6]. These manifestations persist across the lifespan and substantially affect adaptive functioning and quality of life.

Beyond general intellectual impairment, PWS is associated with a distinct cognitive phenotype involving deficits in executive function, working memory, visuospatial processing, and cognitive control [7,8]. Neuropsychological studies have consistently reported impairments in attention, planning, cognitive estimation, inhibitory control, and cognitive flexibility [9,10]. These alterations may contribute to characteristic behavioral features of PWS, including compulsivity, perseveration, impaired set shifting, and difficulties with self-regulation [9,11,12]. Furthermore, psychiatric comorbidities, emotional dysregulation, and variable illness awareness (insight) represent critical components of the adult PWS behavioral spectrum that remain understudied in relation to brain structure. Because these cognitive and behavioral functions depend on the coordinated activity of distributed neural systems, characterizing the neural substrates of PWS requires approaches that extend beyond measures of global intellectual functioning or isolated regional abnormalities.

The distributed nature of this cognitive and behavioral phenotype is consistent with the multisystem effects of PWS. Several genes within the PWS critical region are involved in broad neurodevelopmental, synaptic, neuronal differentiation, and hypothalamic processes, which may influence brain development and function across multiple neural systems rather than producing a focal structural abnormality [13–18]. Consistent with this view, neuroimaging studies have identified structural and functional alterations across distributed cortical and subcortical regions in individuals with PWS. From a systems neuroscience perspective, these alterations may affect the development and organization of large-scale neural networks involved in higher-order cognition, behavioral regulation, and homeostatic control. Characterizing brain architecture at the network level may therefore help link the neurobiological basis of PWS to its complex cognitive and behavioral phenotype.

Previous structural neuroimaging studies in PWS have primarily used voxel-based volumetric methods and reported widespread reductions in gray matter (GM) volume across prefrontal, limbic, subcortical, and cerebellar regions [19–24]. However, volumetric measures provide limited information about the architecture of the cortical mantle. Surface-based morphometry, particularly cortical thickness (CT), offers a complementary and spatially precise characterization of cortical structure. Despite its relevance, CT has not yet been systematically investigated in adults with PWS. Furthermore, previous structural studies have paid limited attention to the organization of cortical abnormalities within large-scale brain networks. Resting-state fMRI (rs-fMRI) studies have defined large-scale functional networks supporting executive control, memory, language, and sensorimotor processing [25,26]. Whether cortical abnormalities in PWS are preferentially distributed across these functionally defined networks remains unknown. Examining cortical architecture within a network framework may therefore provide a more direct link between cortical morphology and the syndrome’s multidimensional clinical phenotype.

PWS also disrupts hypothalamic–pituitary function, with growth hormone (GH) deficiency being a common endocrine abnormality [6]. GH treatment improves growth, body composition, and developmental outcomes in children and is generally safe and beneficial for body composition in adults [27,28]. Its potential neurodevelopmental effects, particularly when initiated early [29], suggest that adult brain structure may reflect not only the underlying genetic disorder but also associated endocrine dysfunction and its treatment.

The present study aimed to characterize CT in adults with PWS by comparing 23 individuals with PWS with 22 age- and sex-matched healthy controls. We examined group differences in CT both at the regional level and by mapping significant cortical alterations onto large-scale functional networks, and assessed associations between CT and demographic, cognitive, anthropometric, endocrine, and functional measures. Based on previous volumetric findings and the cognitive–behavioral phenotype of PWS, we hypothesized that individuals with PWS would show reduced CT in distributed cortical regions implicated in executive, attentional, and sensorimotor functions. This surface-based approach extends previous volumetric findings by examining cortical architecture within a network-level framework and relating cortical morphology to the multidimensional clinical phenotype of PWS.

## Methods and Materials

### Subjects and Study

Participants were drawn from a cohort previously described elsewhere, in which the effects of one year of rhGH treatment on body composition, motor function and brain functional activation were reported [27]. The present study analyzed baseline structural MRI data from this cohort to address a distinct research question and included measures that have not been previously reported.

From that cohort, our study included a total of 46 participants: 23 adults with PWS and 23 healthy controls matched for age and sex. One healthy control was excluded from the analysis due to excessive head motion during MRI acquisition; thus, analyses were performed with 23 adults with PWS and 22 healthy controls (45 participants in total). Descriptive characteristics of both study groups are shown in Table 1.

**Table 1.** Descriptive characteristics of the two participant cohorts. Continuous variables are presented as mean ± standard deviation (SD) and categorical variables as absolute frequencies. Psychiatric diseases were defined as any current psychiatric comorbidity. Psychotropic medications included antipsychotics, mood stabilizers, benzodiazepines and/or antidepressants. GH therapy during childhood refers to previous treatment and was not ongoing at the time of assessment. Abbreviations: BMI, body mass index; eTIV, estimated total intracranial volume; F, female; GH, growth hormone; ID, imprinting defect; L, left; M, male; NA: not applicable; PWS, Prader– Willi syndrome; R, right; SD, standard deviation; UPD, maternal uniparental disomy.

|  | Group: mean ± SD |  |
|  | PWS (n = 23) | Control (n = 22) |
|---|---|---|
| Age, yr | 30.6 ± 10.1 | 29.8 ± 9.2 |
| Sex, M/F | 11/12 | 10/12 |
| BMI, kg/m² | 34.9 ± 7.4 | 22.0 ± 1.8 |
| Body fat, % | 54.3 ± 7.1 | 32.9 ± 7.1 |
| eTIV, cm³ | 1447 ± 109 | 1590 ± 120 |
| Handedness, R/L | 16/7 | 18/4 |
| Genetic diagnosis, n |  |  |
| *Type 1 Deletion* | 6 | NA |
| *Type 2 Deletion* | 8 | NA |
| *Atypical Deletion* | 2 | NA |
| *UPD* | 4 | NA |
| *ID* | 3 | NA |
| Comorbidities |  |  |
| *Type 2 Diabetes Mellitus, yes/no* | 5/18 | 0/22 |
| *Hypertension, yes/no* | 2/21 | 0/22 |
| *Dyslipidemia, yes/no* | 5/18 | 0/22 |
| *Hypogonadism, yes/no* | 23/0 | 0/22 |
| *Hypothyroidism, yes/no* | 3/20 | 0/22 |
| *Obstructive Sleep Apnea, yes/no* | 15/8 | 0/22 |
| *Psychiatric Diseases, yes/no* | 17/6 | 0/22 |
| Pharmacological Treatments |  |  |
| *GH therapy during childhood, yes/no* | 13/10 | 0/22 |
| *Psychotropics, yes/no* | 14/9 | 0/22 |
| *Sex Hormone Replacement Therapy, yes/no* | 12/11 | 0/22 |

The study protocol received approval from the local ethics committee (Institutional Ethics Committee of Institut d’Investigació i Innovació Parc Taulí, Sabadell, Barcelona; ref. END-GH-2017) and adhered to the principles of the Declaration of Helsinki. Adults with PWS and their legal guardians were fully informed about the study. Participants provided assent, and their legal guardians provided written informed consent. Control participants also provided written informed consent.

### MRI acquisition

Structural MRI scans were acquired on a 3.0 T Philips Achieva system (Philips Healthcare, Best, The Netherlands; RRID:SCR_008656) with an eight-channel phased-array head coil. T1-weighted anatomical images were collected using a 3D fast spoiled gradient inversion-recovery sequence, acquiring 160 contiguous slices (repetition time: 8.2 ms, echo time: 3.792 ms, flip angle: 8°, field of view: 25.6 cm, matrix: 256 × 256 pixels, slice thickness: 1 mm).

### Cortical thickness analysis

Cortical reconstruction and volumetric segmentation analysis of participants’ structural MR images was run using FreeSurfer brain imaging software package version 8.0.0, which is documented and freely available for download online at http://surfer.nmr.mgh.harvard.edu. Prior to the analysis all T1 images were visually examined for common MRI artifacts such as ghosting and ring-motion. All participants’ MR images were processed with the automatic FreeSurfer “recon-all” pipeline. The technical details of these procedures are described in prior publications [30–32] and available at (https://surfer.nmr.mgh.harvard.edu/fswiki/FreeSurferMethodsCitation). Briefly, this processing includes motion correction, removal of non-brain tissue using a hybrid watershed/surface deformation procedure, automated Talairach transformation, segmentation of the subcortical white matter and deep gray matter volumetric structures, intensity normalization, tessellation of the gray matter white matter boundary, automated topology correction, and surface deformation following intensity gradients to optimally place the gray/white and gray/cerebrospinal fluid borders at the location where the greatest shift in intensity defines the transition to the other tissue class.

Once the cortical models were complete, reconstructed and registered individual CT maps were smoothed using Gaussian kernels of 10 and 15 mm full-width half maximum and subsequently used in statistical analyses. Procedures for the measurement of CT have been validated against histological analysis [33] and manual measurements [34,35]. FreeSurfer morphometric procedures have been demonstrated to show good test-retest reliability across scanner manufacturers and across field strengths [36,37].

### Cortical parcellation

CT analyses typically involve parcellating the cortical ribbon into anatomically defined regions of interest (ROIs) based on a standardized template. To achieve this, the reconstructed cortical surface is inflated and registered to a spherical atlas using individual gyral and sulcal folding patterns [30,32]. In the present study, cortical parcellation was performed using the Destrieux atlas [38], which subdivides each cerebral hemisphere into 74 anatomically defined ROIs based on sulco-gyral anatomy. The Destrieux atlas was selected over coarser parcellation schemes, such as the Desikan–Killiany atlas, to provide higher anatomical resolution and increased sensitivity to localized CT differences, which is particularly relevant for detecting subtle and spatially specific alterations in clinical populations.

To examine CT within the framework of large-scale cognitive systems, we employed a predefined cortical network atlas. Specifically, we used the DU15NET-Consensus atlas (Figure 1), a recently released parcellation in which large-scale functional networks (resting state networks; RSN) were derived from resting-state fMRI data collected in intensively sampled participants [39]. This consensus atlas delineates 15 cortical networks and was designed for use in group-level analyses. It represents an update to the widely used Yeo et al. [40] network parcellations, incorporating refinements that reflect current understanding of human cerebral network organization [39].

**FIGURE 1.**
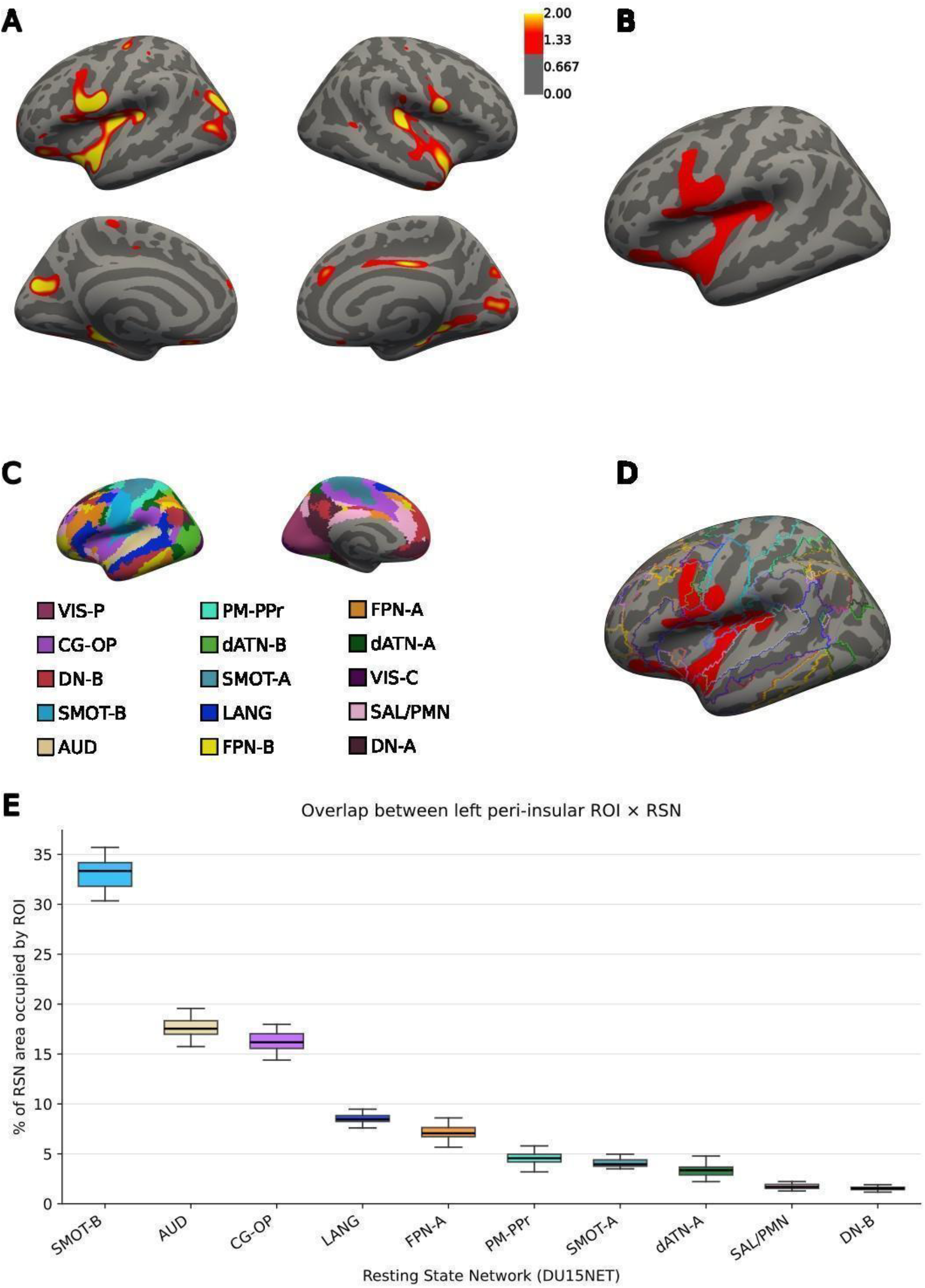
CT comparison between PWS patients and controls. A) Uncorrected comparison of CT at 15 mm FWHM, with a threshold 1-2, and directionality C > PWS. B) Peri-insular region showing significantly reduced CT in the PWS group compared with controls after cluster-wise correction. Cluster size was 4518.5 mm^2^, with a p-value of 0.0486. C) Atlas of DU15NET resting state networks. Abbreviations: AUD, Auditory; CG-OP, Cingulo-Opercular; dATN-A, Dorsal Attention-A; dATN-B, Dorsal Attention-B; DN-A, Default Network-A; DN-B, Default Network-B; FPN-A, Frontoparietal Network-A; FPN-B, Frontoparietal Network-B; LANG, Language; PM-PPr, Premotor-Posterior Parietal Rostral; SAL/PMN, Salience/Parietal Memory Network; SMOT-A, Somatomotor-A; SMOT-B, Somatomotor-B; VIS-C, Visual Central; VIS-P, Visual Peripheral. D) Overlap between the region showing significantly reduced CT and the different RSNs. E) Boxplots show the percentage of vertices within each resting-state network (RSN) that overlapped with the region of interest (ROI). Percentages were normalized by the total number of vertices in each RSN. Only RSNs with non-zero overlap with the ROI are shown.

### Behavioral and Clinical Variables

Participants from the PWS group were extensively phenotyped across multiple domains. Demographic variables included age and sex. Anthropometric and physical-function measures included body mass index (BMI), body fat percentage, handgrip strength assessed with a Jamar dynamometer [41], and balance and functional mobility with Berg Balance Scale [42] and Timed Up and Go test [43]. Intellectual functioning was assessed using the Kaufman Brief Intelligence Test (K-BIT) [44], whereas global functioning was assessed using the Global Assessment of Functioning (GAF) [45]. Awareness of illness and syndrome characterization were evaluated using PWS-adapted scales [46,47]. Patient consciousness of the syndrome (PCS) and syndrome severity (SS) were assessed using Likert-type visual analogue scales (0 to 10), and patient total awareness of illness (PTAI) was evaluated using a multidimensional insight scale adapted for PWS, where higher scores indicate lower insight and attribution. Psychological functioning was assessed using the Strengths and Difficulties Questionnaire (SDQ) [48,49] administered to primary caregivers to evaluate emotional and behavioral difficulties across five subscales (emotional symptoms, conduct problems, hyperactivity/inattention, peer relationship problems, and prosocial behavior). Hormonal profiles were obtained from peripheral blood samples and included insulin-like growth factor (IGF), leptin, and kisspeptin levels. GH treatment during childhood was also registered. For the control group, only anthropometric measures were acquired. Descriptive data for all variables are available in Table 2, except for BMI and body fat percentage (Table 1).

**Table 2.**
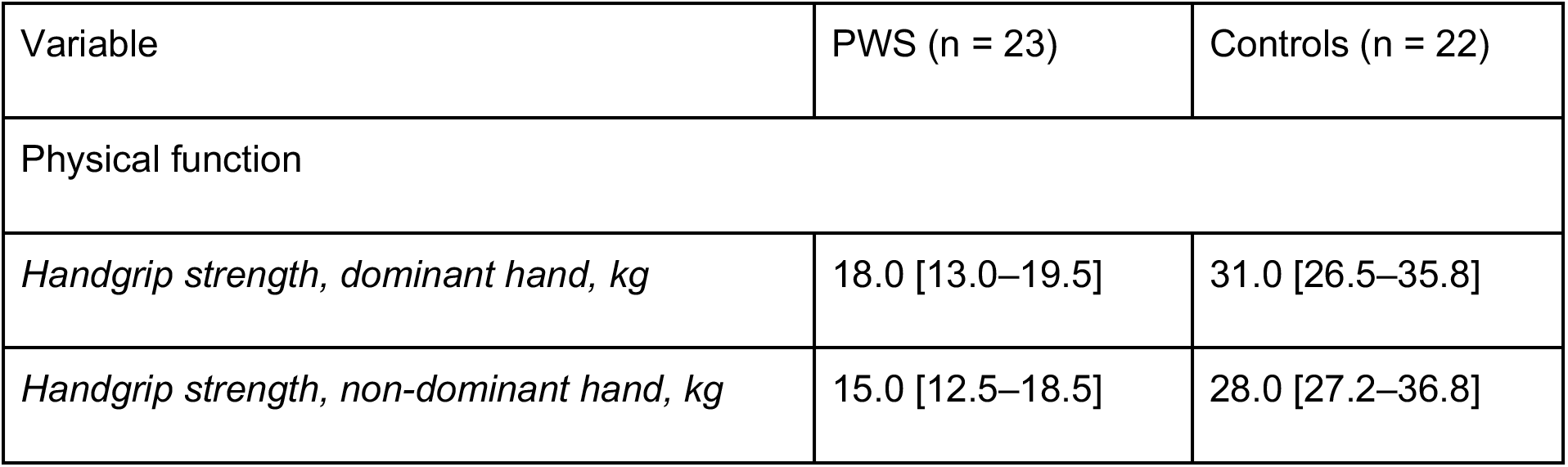
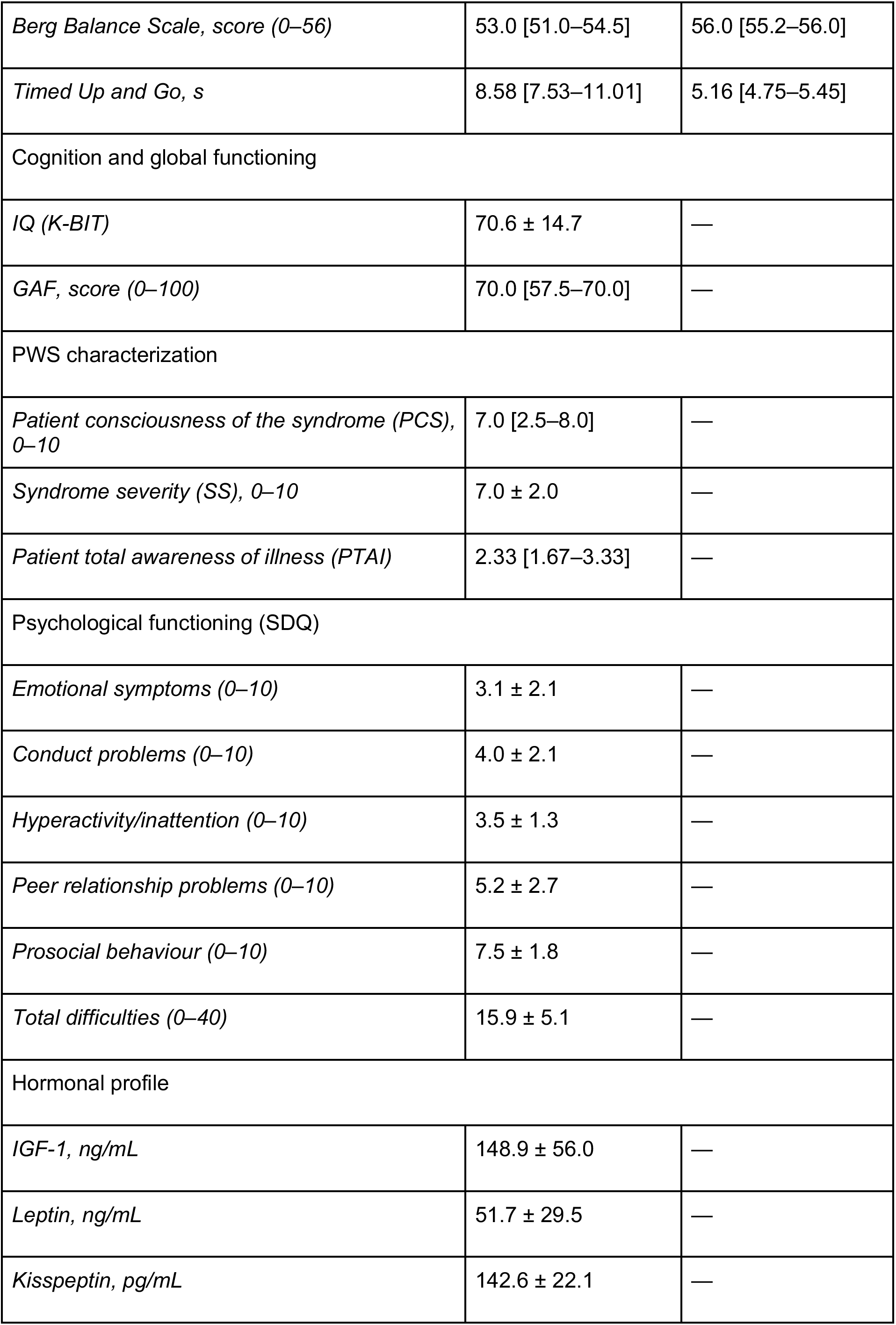
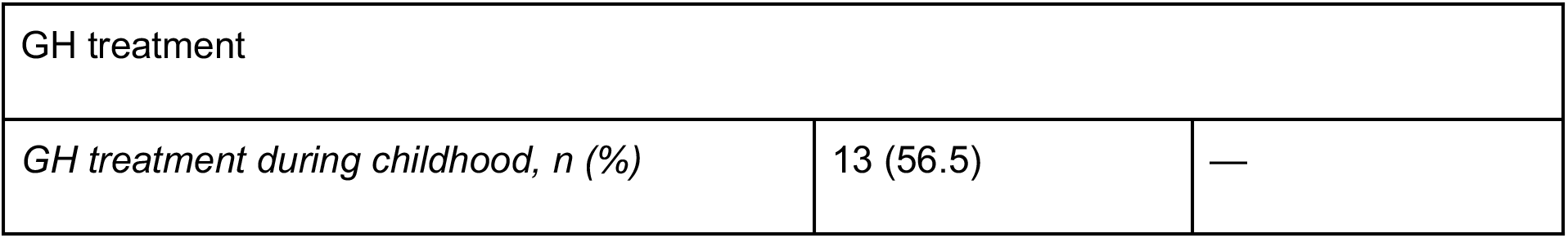
Physical, cognitive, behavioral and hormonal characteristics of the participants. Values are mean ± SD for normally distributed variables and median [IQR] for non-normally distributed variables. Categorical variables are n (%). Abbreviations: GAF, Global Assessment of Functioning; GH, growth hormone; IGF-1, insulin-like growth factor 1; K-BIT, Kaufman Brief Intelligence Test; PCS, patient consciousness of the syndrome; PTAI, patient total awareness of illness (higher scores indicate worse insight and attribution); PWS, Prader–Willi syndrome; SDQ, Strengths and Difficulties Questionnaire; SS, syndrome severity.

### Statistical analysis

Group differences in CT were assessed by comparing reconstructed and registered individual CT maps across the entire cortical surface. Surface-based smoothing was applied using Gaussian kernels with full widths at half maximum (FWHM) of 10 and 15 mm. Vertex-wise group comparisons between individuals with PWS and healthy controls were performed using a general linear model (GLM) with a different-offset, different-slope design. Age, sex, and estimated total intracranial volume (eTIV) [50] were included as nuisance covariates. Surface-based statistical analyses were conducted using standard routines implemented in the FreeSurfer software suite, including mris_preproc, mri_glmfit, and mri_glmfit-sim. Correction for multiple comparisons was performed using cluster-wise inference based on Monte Carlo simulations with 5,000 iterations applying a threshold of 1.3 (p=0.05) one-tailed. Following cluster-wise correction, post hoc analyses were performed on the resulting significant cortical regions.

Also, group differences in CT were assessed at regional anatomical level using the Destrieux atlas. Mean CT values for each region were extracted using the aparcstats2table command and analyzed using analysis of covariance (ANCOVA), with age, sex, and estimated total intracranial volume (eTIV) included as nuisance covariates. P-values were adjusted (q) for multiple comparisons.

For each participant with PWS, we characterized the distribution of the significant region of interest (ROI) across the large-scale RSNs defined by the DU15NET-Consensus atlas. Specifically, for each RSN, overlap was quantified as the percentage of RSN vertices included within the ROI, thereby normalizing the ROI–RSN overlap by the total number of vertices in that RSN. Participant-level values were subsequently averaged and reported at the group level, as follows:

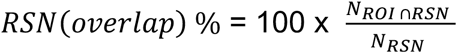

To investigate the relationship between CT and behavioral and clinical measures, Spearman rank correlation analyses were performed to examine associations between participants’ behavioral and clinical variables and mean CT values extracted from the potential ROIs identified in FreeSurfer comparisons. Correlations were computed separately within each group, and between-group differences in correlation coefficients were assessed using Fisher’s r-to-z transformation. Between-group differences were possible only for anthropometric measures. P-values were adjusted (q) for multiple comparisons within groups of variables (anthropometric, cognitive, PWS characterization, hormonal and psychological).

For differences within the PWS group between individuals who received GH replacement therapy during childhood and those who did not, a Welch t test was performed to compare mean CT of the potential ROIs identified during FreeSurfer comparisons.

## Results

At the pre–cluster-correction level (15 mm FWHM), group differences were bilateral and approximately symmetric, involving the inferior temporal cortex, orbitofrontal cortex, visual areas, and motor cortex (Figure 1A). After cluster-wise correction at the same smoothing level, CT differences were confined to the left hemisphere, with a single cluster (4518.5 mm^2^) spanning the inferior precentral gyrus, insula, inferior temporal cortex, and orbitofrontal cortex (Figure 1B).

At the ROI level, ANCOVA using the Destrieux atlas revealed regional differences in CT that did not survive multiple-comparison correction (Table S1); however, the pattern was consistent with whole-brain findings, with reduced CT in the insula, temporal, and orbitofrontal regions in the PWS group.

To contextualize the spatial distribution of the cluster-corrected findings, we examined their overlap with large-scale functional networks defined by the DU15NET-Consensus atlas (Figure 1C). The highest ROI–RSN overlap percentages were observed for the somatomotor-B (SMOT-B; 33%), auditory (AUD; 18%), and cingulo-opercular (CG-OP; 17%) networks, followed by the language (LANG; approximately 8%) and frontoparietal-A (FPN-A; approximately 7%) networks.

At the behavioral and clinical level, we examined associations between mean CT within the identified ROI and anthropometric, cognitive, syndrome characterization, and hormonal measures. Fisher’s r-to-z comparisons revealed significant between-group differences in the association between BMI and mean CT (mm) (z = 3.23, q = 0.007). Specifically, within the PWS group, BMI was significantly negatively correlated with mean CT (*ρ* = -0.66, q = 0.003), whereas the control group showed a non-significant positive correlation (*ρ* = 0.23). No other anthropometric associations were significant (Figure 2).

**FIGURE 2.**
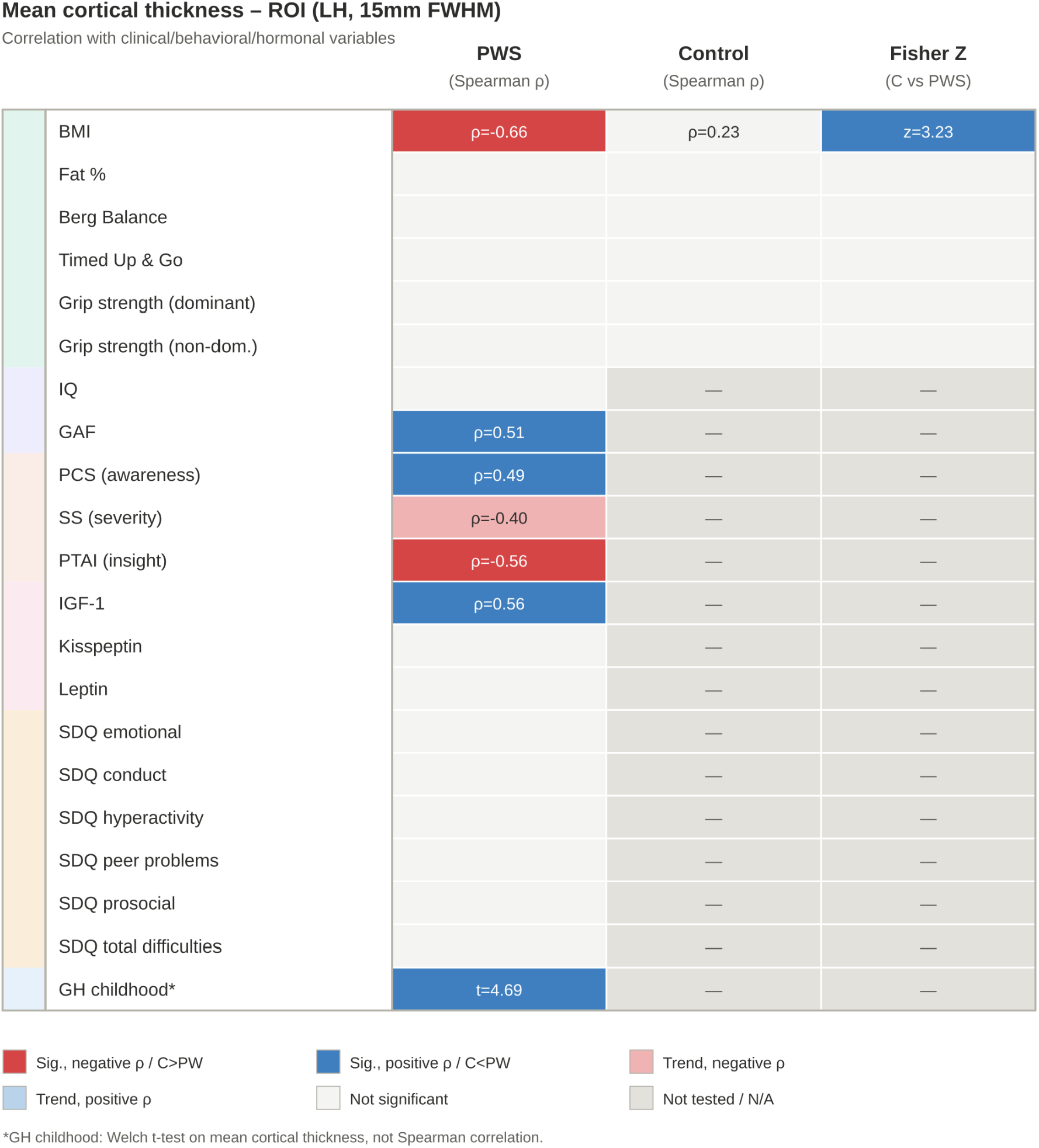
Correlations between mean CT within the significant left-hemisphere ROI (15 mm FWHM, cluster-corrected) and clinical, cognitive, behavioral, hormonal, and GH treatment-related variables. Cell values represent Spearman’s rho, computed separately within the PWS and control groups; the third column shows the Z statistic from Fisher’s r-to-z test comparing correlation coefficients between controls and participants with PWS, calculated as *Z_control_*−*Z_P_*_W*S*_. Positive values (red) indicate stronger correlations in controls than in PWS, whereas negative values (blue) indicate stronger correlations in PWS than in controls. Saturated colors denote statistically significant associations (q < 0.05, FDR-corrected within group of variables); pale shades denote trend-level associations (q < 0.10). Light gray cells indicate non-significant associations; dark gray cells indicate variables not tested or not acquired for that group (only anthropometric measures were collected in controls). The GH childhood (yes/no) row reflects a Welch’s t-test comparing mean CT between PWS participants with and without growth hormone treatment during childhood, rather than a Spearman correlation. Abbreviations: BMI, body mass index; GAF, Global Assessment of Functioning; PCS, patient consciousness of the syndrome; SS, syndrome severity; PTAI, patient total awareness of the illness; IGF-1, insulin-like growth factor I; SDQ, Strengths and Difficulties Questionnaire.

In terms of cognitive functioning, mean CT was positively correlated with GAF scores in the PWS group, surviving correction (*ρ* = 0.51, q = 0.026), with no significant association observed for IQ (Figure 2).

Regarding syndrome characterization, CT was positively correlated with the PCS (*ρ* = 0.49, q = 0.045) and negatively correlated with the PTAI (*ρ* = -0.56, q = 0.045). A trend-level negative association was observed with SS (*ρ* = -0.40, q = 0.06) (Figure 2).

Finally, mean CT showed a strong positive association IGF-1 levels (*ρ* = 0.56, q = 0.01), and was significantly greater in participants with a history of previous GH replacement therapy during childhood (t = 4.69, p < 0.001). No significant associations were observed for other hormonal or psychological variables (Figure 2).

## Discussion

This study provides a surface-based analysis of CT in adults with PWS, extending previous volumetric findings by examining cortical morphology at the surface level and within a large-scale network framework. We identified a cluster of reduced CT in the left hemisphere involving the inferior precentral gyrus, insula, inferior temporal cortex, and orbitofrontal cortex. A spatially similar pattern was also observed in the right hemisphere at an uncorrected threshold, but did not survive correction for multiple comparisons. The significant cluster showed anatomical overlap with the somatomotor, auditory, cingulo-opercular, language, and frontoparietal networks. Within this region, CT was associated with BMI, global functioning, selected syndrome-related measures, IGF-1 concentrations, and a history of GH treatment during childhood. Together, these findings suggest that cortical morphology in adults with PWS is altered across distributed regions involved in sensorimotor, cognitive, and behavioral processes and may be related to both metabolic and endocrine characteristics of the syndrome.

### Comparison with previous structural neuroimaging findings

The anatomical distribution of CT reductions is broadly consistent with previous voxel-based morphometry (VBM) studies reporting reduced gray matter (GM) volume in overlapping regions in PWS, particularly the orbitofrontal cortex [19,20]. Honea et al. [20], also reported reductions in prefrontal and orbitofrontal cortices, temporal regions, the hippocampus, and parahippocampal gyrus, whereas Caixàs et al. [21] identified reduced GM volume in a small portion of the left caudate nucleus and cerebellum.

The present findings extend these observations by providing greater anatomical specificity at the cortical level. Whereas VBM measures regional GM volume, surface-based morphometry allows CT to be examined independently of other components contributing to regional volume. The overlap between our CT findings and previously reported volumetric abnormalities raises the possibility that reduced CT contributes to some of these findings, although methodological differences between VBM and surface-based approaches must also be considered.

Our findings also differ from those of Manning et al. [22], who reported both increases and decreases in GM volume in a younger cohort of adults with PWS, including increased GM volume in medial prefrontal and cingulate regions. These discrepancies may stem from differences in sample age, MRI acquisition, and analysis protocols, as well as age-related brain maturation and the cumulative effects of obesity and endocrine abnormalities. Direct comparisons across studies should therefore be interpreted cautiously.

### Hemispheric distribution and network-level interpretation

The predominance of significant CT differences in the left hemisphere, despite a broadly bilateral pattern at the uncorrected threshold, is noteworthy given that previous neuroimaging studies of PWS have often reported bilateral or multiregional structural abnormalities [22,51]. This asymmetry should not be interpreted as evidence of a truly lateralized PWS phenotype, as the modest sample size may have limited power to detect bilateral effects after correction. Larger samples will be needed to determine whether the left-lateralized pattern is reproducible.

The significant cluster overlapped predominantly with the somatomotor, auditory, and cingulo-opercular networks, with additional involvement of language and frontoparietal systems. This distribution extends beyond the limbic and reward-related regions traditionally emphasized in PWS and suggests involvement of systems supporting sensorimotor integration, attention, executive control, and language. Such a distributed pattern is consistent with the multidimensional developmental phenotype of the syndrome [51].

The involvement of the somatomotor network, together with the extension of the cluster into the insula and orbitofrontal cortex, may be relevant not only to motor abnormalities in PWS [52] but also to eating behavior. Indeed, the inferior precentral gyrus, the anterior insula and adjacent frontal operculum, and the orbitofrontal cortex have been implicated in orofacial motor control, gustatory processing, and the encoding of food reward and satiety [53]. This interpretation is consistent with functional imaging studies showing altered responses to food stimuli in the insula and orbitofrontal cortex in PWS [54,55]. However, hyperphagia-specific measures were not available in our study, and the relevance of this cortical finding to eating behavior remains to be directly tested.

Similarly, the involvement of the cingulo-opercular network, which has been implicated in sustained attention and cognitive control, may be relevant to the attentional and executive difficulties reported in PWS [7]. Anatomical overlap with auditory and language-related regions may likewise be relevant to the language and communication difficulties described in the syndrome [56]. However, these interpretations remain hypothesis-generating, as anatomical overlap with functional network templates does not by itself demonstrate functional dysfunction within these networks. Future multimodal studies combining cortical morphology with functional connectivity will be important for determining whether the structural alterations identified here are accompanied by altered network function.

### Associations with BMI and clinical functioning

Within the PWS group, BMI was negatively associated with CT (*ρ* = −0.66, q = 0.003), while no significant association was found in the control group. The difference between correlations was significant (z = 3.23, q = 0.007), although this comparison should be interpreted cautiously, as the BMI range in controls was much narrower, which limits the ability to detect an association in that group. The negative association in PWS is consistent with previous studies in pediatric populations [57] and adults without PWS [58,59], which have reported inverse associations between BMI and CT, particularly in prefrontal regions.

Although chronic obesity may affect brain structure through metabolic and inflammatory pathways, CT was not associated with body fat percentage (ρ = 0.12, p = 0.58) or leptin concentrations (ρ = 0.10, p = 0.66). These findings provide little support for a simple relationship between CT and adiposity-related biomarkers. BMI may instead reflect broader features of the PWS phenotype, including GH/IGF-1 status, hyperphagia, lifelong environmental control of food intake, and cumulative obesity exposure. Participants treated with GH during childhood had lower BMI than untreated participants (32.3 ± 7.2 vs. 38.3 ± 6.6 kg/m², p = 0.03), although BMI was not significantly associated with IGF-1 concentrations (ρ = −0.32, p = 0.14). Longitudinal studies are therefore needed to determine whether changes in body composition are accompanied by changes in cortical morphology.

CT was also positively associated with GAF scores (*ρ* = 0.51, q = 0.026) suggesting an association between cortical morphology and clinically meaningful aspects of functioning. Interestingly, cortical alterations were not significantly associated with general IQ, but specifically with broader adaptive and psychosocial functioning (GAF) as well as syndrome-specific measures (PCS, PTAI, SS). This dissociation suggests that cortical morphology in adults with PWS tracks dimensional features of daily-life adaptation and self-referential cognitive processing rather than static global intelligence. Regarding psychological functioning evaluated via the SDQ, no direct correlations were found with cortical thickness in our cohort. While the SDQ provides valuable dimensional screening for emotional and behavioral difficulties in daily life, these broad psychological measures likely capture complex behavioral phenotypes and systemic dynamics that extend beyond regional cortical morphology, relying instead on widespread multi-system networks and psychosocial factors. Despite the exploratory nature of these associations, results suggest that cortical morphology captures dimensional aspects of phenotypic variability beyond global cognitive metrics.

### Association with the GH/IGF-1 axis

The positive association between CT and circulating IGF-1 levels (*ρ* = 0.56, q = 0.01), together with the difference in CT between participants with and without a history of childhood GH therapy (t = 4.69, p < 0.001), suggest that the GH–IGF-1 axis may be associated with cortical maturation in this population. GH treatment has beneficial effects on body composition, linear growth, and aspects of cognitive development in PWS, with earlier treatment associated with better developmental outcomes [60–62]. Benefits have also been reported when treatment is initiated during adulthood [27].

The GH/IGF-1 axis contributes to neurogenesis, synaptic development, and myelination [63–65]. Children with isolated GH deficiency have been reported to show reductions in GM volume and cortical surface area compared with controls, with positive associations between IGF-1 concentrations and cortical morphometric measures [63]. In the general population, higher IGF-1 concentrations have likewise been associated with greater total brain and hippocampal volumes [66,67]. These observations provide a plausible biological framework for the positive association between GH/IGF-1 signaling and CT observed in PWS.

However, the cross-sectional association between a history of childhood GH therapy and adult CT may reflect direct effects of GH/IGF-1 signaling, as well as indirect effects mediated through body composition or metabolic health, or differences between individuals who did and did not receive treatment. Treatment history was not randomly assigned, and factors influencing the treatment initiation may themselves be related to long-term neurodevelopmental outcomes. Therefore, longitudinal studies with repeated measures of GH exposure, IGF-1, body composition, and brain morphology will be needed to clarify these relationships.

### Limitations and future directions

This study should be interpreted considering its modest sample size and cross-sectional design. The sample size is comparable to or larger than that of many previous neuroimaging studies in PWS and enabled the detection of significant cortical and clinical associations [22,23,51]. Nevertheless, larger multicenter studies will be useful for confirming the hemispheric distribution of the findings and examining potential genetic subtype effects. Longitudinal studies will also be important for clarifying the temporal relationship among CT, BMI, IGF-1 levels, and GH treatment history. administered to primary caregivers to evaluate emotional and behavioral difficulties across five subscales (emotional symptoms, conduct problems, hyperactivity/inattention, peer relationship problems, and prosocial behavior).

## Conclusions

Overall, these findings provide evidence of altered CT in adults with PWS and extend previous volumetric observations by showing that structural abnormalities are distributed across cortical regions mapping onto multiple functional systems. Associations of CT with BMI, global functioning, syndrome-related measures, GH treatment history, and IGF-1 levels indicate that cortical morphology reflects clinically relevant dimensions of the PWS phenotype. These findings suggest an interplay between underlying genetic vulnerability and modifiable endocrine-metabolic factors, pointing to a potential neuroprotective role for GH/IGF-1 signaling. Although these findings require replication in larger and longitudinal cohorts, they are consistent with the possibility that genetic, metabolic, endocrine, and neurodevelopmental factors jointly influence cortical morphology across multiple large-scale brain networks in PWS.

## Acknowledgements

CPG acknowledges his AI4S fellowship within the “Generación D” initiative by Red.es, Ministerio para la Transformación Digital y de la Función Pública, for talent attraction (C005/24-ED CV1), funded by NextGenerationEU through PRTR.

We are indebted to all participants and their families who collaborated in all the studies involved in the present manuscript.

## Data Availability

Data Availability Statement: Individual-level structural MRI and clinical data are not publicly available because of participant privacy and ethical restrictions but may be made available upon reasonable request, subject to institutional and ethical approval.

Declaration of generative AI and AI-assisted technologies in the manuscript preparation process.

During the preparation of this work the author(s) used Claude; Anthropic for (1) grammar checking and English-language correction during the preparation of the manuscript and (2) code debugging. After using this tool/service, the author(s) reviewed and edited the content as needed and take(s) full responsibility for the content of the published article.

## Author Contributions

**Alex M-A:** Conceptualization, Methodology, Formal analysis, Investigation, Data Curation, Writing – Original Draft, Writing – Review & Editing, Software, Project Administration, Visualization, Validation.

**Laura B:** Data Curation, Methodology, Formal analysis, Investigation, Writing – Review & Editing, Resources, Validation, Supervision.

**Laia C:** Conceptualization, Investigation, Methodology, Formal analysis, Validation, Writing – review and editing.

**Olga G-P:** Conceptualization, Investigation, Methodology, Formal analysis, Validation, Writing – review and editing.

**Jesús C:** Methodology, Investigation, Validation, Writing – review and editing.

**Jesús P:** Conceptualization, Data Curation, Methodology, Formal analysis, Investigation, Supervision, Writing – review and editing.

**Joan D:** Conceptualization, Data Curation, Methodology, Formal analysis, Investigation, Supervision, Writing – review and editing.

**Susanna E-C**: Methodology, Investigation, Writing – Review & Editing.

**Rocío P:** Methodology, Investigation, Writing – Review & Editing.

**Assumpta C**: Conceptualization, Methodology, Formal analysis, Investigation, Writing – Original Draft, Writing – Review & Editing, Resources, Funding Acquisition, Project Administration, Validation, Supervision.

**Cleofé PG:** Conceptualization, Methodology, Formal analysis, Investigation, Data Curation, Writing – Original Draft, Writing – Review & Editing, Software, Funding Acquisition, Project Administration, Visualization, Validation, Supervision. All authors reviewed and approved the final manuscript.

---

**Supplementary Table 1.**
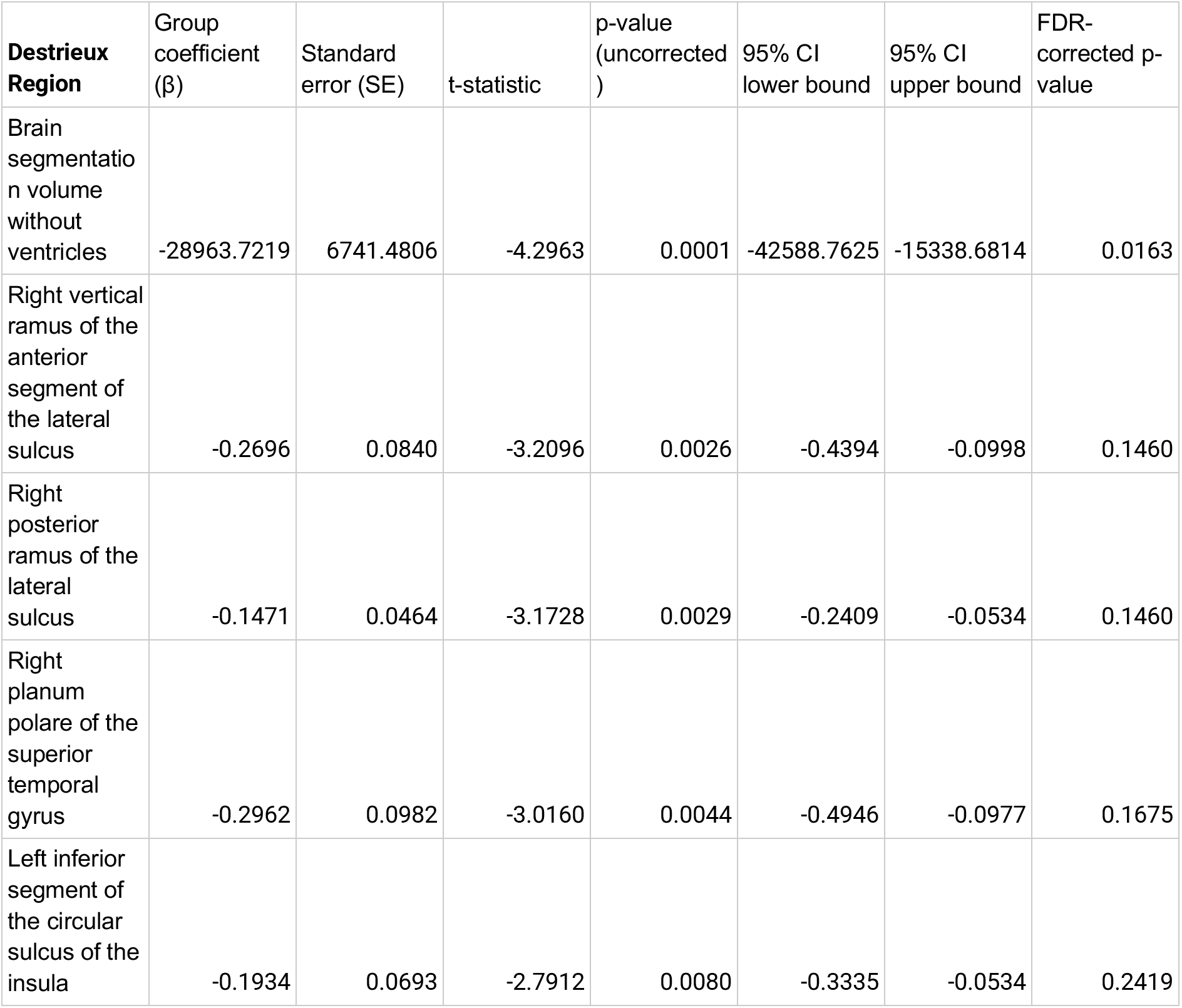
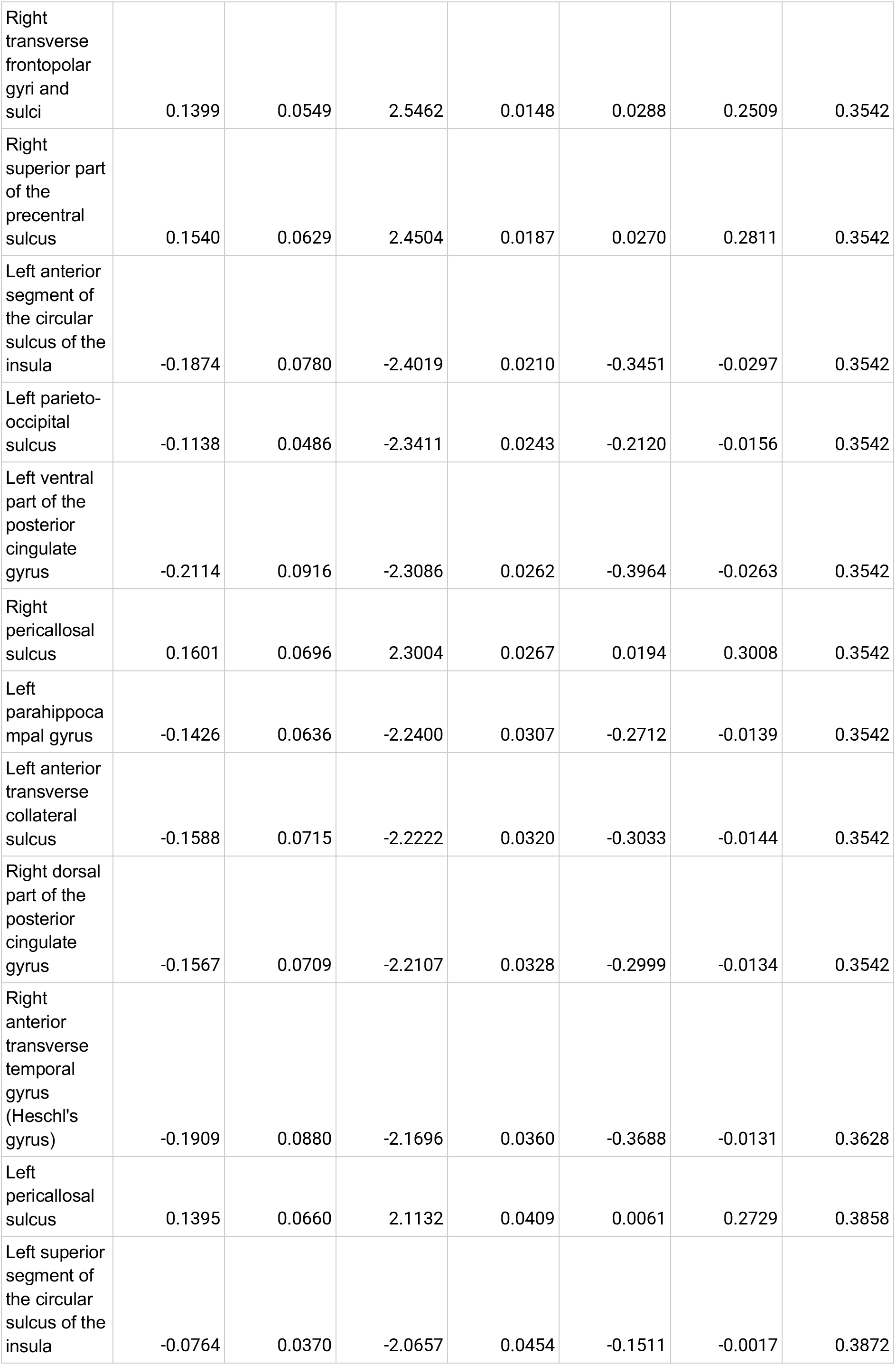
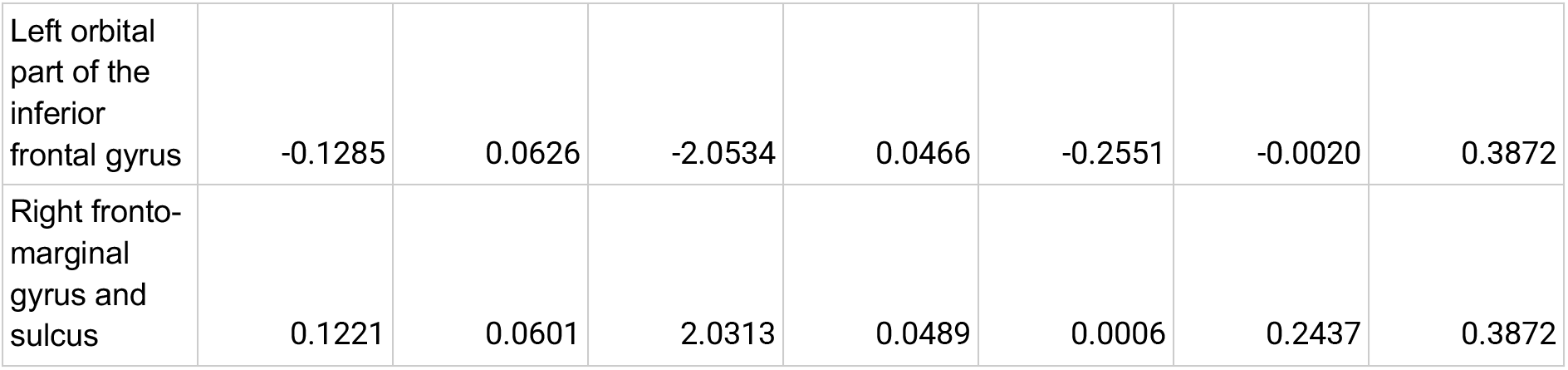
Regional cortical differences between adults with Prader-Willi syndrome (PWS, n = 23) and healthy controls (n = 22) in the Destrieux atlas (aparc.a2009s). Group differences were tested with ANCOVA for each of the 151 variables, adjusting for age, sex and estimated total intracranial volume (eTIV). Only variables with an uncorrected p < 0.05 are shown. The false discovery rate (FDR) correction was applied across all 151 variables using the Benjamini–Hochberg procedure. β is the adjusted group coefficient ([PWS − controls]; negative values indicate lower values in PWS), expressed in mm. SE: standard error; CI: confidence interval; FDR: false discovery rate.

| **Destrieux Region** | Group coefficient (β) | Standard error (SE) | t-statistic | p-value (uncorrected) | 95% CI lower bound | 95% CI upper bound | FDR-corrected p-value |
|---|---|---|---|---|---|---|---|
| Brain segmentation volume without ventricles | -28963.7219 | 6741.4806 | -4.2963 | 0.0001 | -42588.7625 | -15338.6814 | 0.0163 |
| Right vertical ramus of the anterior segment of the lateral sulcus | -0.2696 | 0.0840 | -3.2096 | 0.0026 | -0.4394 | -0.0998 | 0.1460 |
| Right posterior ramus of the lateral sulcus | -0.1471 | 0.0464 | -3.1728 | 0.0029 | -0.2409 | -0.0534 | 0.1460 |
| Right planum polare of the superior temporal gyrus | -0.2962 | 0.0982 | -3.0160 | 0.0044 | -0.4946 | -0.0977 | 0.1675 |
| Left inferior segment of the circular sulcus of the insula | -0.1934 | 0.0693 | -2.7912 | 0.0080 | -0.3335 | -0.0534 | 0.2419 |
| Right transverse frontopolar gyri and sulci | 0.1399 | 0.0549 | 2.5462 | 0.0148 | 0.0288 | 0.2509 | 0.3542 |
| Right superior part of the precentral sulcus | 0.1540 | 0.0629 | 2.4504 | 0.0187 | 0.0270 | 0.2811 | 0.3542 |
| Left anterior segment of the circular sulcus of the insula | -0.1874 | 0.0780 | -2.4019 | 0.0210 | -0.3451 | -0.0297 | 0.3542 |
| Left parieto-occipital sulcus | -0.1138 | 0.0486 | -2.3411 | 0.0243 | -0.2120 | -0.0156 | 0.3542 |
| Left ventral part of the posterior cingulate gyrus | -0.2114 | 0.0916 | -2.3086 | 0.0262 | -0.3964 | -0.0263 | 0.3542 |
| Right pericallosal sulcus | 0.1601 | 0.0696 | 2.3004 | 0.0267 | 0.0194 | 0.3008 | 0.3542 |
| Left parahippocampal gyrus | -0.1426 | 0.0636 | -2.2400 | 0.0307 | -0.2712 | -0.0139 | 0.3542 |
| Left anterior transverse collateral sulcus | -0.1588 | 0.0715 | -2.2222 | 0.0320 | -0.3033 | -0.0144 | 0.3542 |
| Right dorsal part of the posterior cingulate gyrus | -0.1567 | 0.0709 | -2.2107 | 0.0328 | -0.2999 | -0.0134 | 0.3542 |
| Right anterior transverse temporal gyrus (Heschl's gyrus) | -0.1909 | 0.0880 | -2.1696 | 0.0360 | -0.3688 | -0.0131 | 0.3628 |
| Left pericallosal sulcus | 0.1395 | 0.0660 | 2.1132 | 0.0409 | 0.0061 | 0.2729 | 0.3858 |
| Left superior segment of the circular sulcus of the insula | -0.0764 | 0.0370 | -2.0657 | 0.0454 | -0.1511 | -0.0017 | 0.3872 |
| Left orbital part of the inferior frontal gyrus | -0.1285 | 0.0626 | -2.0534 | 0.0466 | -0.2551 | -0.0020 | 0.3872 |
| Right fronto-marginal gyrus and sulcus | 0.1221 | 0.0601 | 2.0313 | 0.0489 | 0.0006 | 0.2437 | 0.3872 |

